# A Bayesian hierarchical framework to integrate dietary exposure and biomarker measurements into aetiological models

**DOI:** 10.1101/2022.03.24.22272838

**Authors:** M. Pittavino, M. Plummer, M. Johansson, P. Ferrari

## Abstract

In nutritional epidemiology, self-reported assessments of dietary exposure are prone to measurement errors, which is responsible for bias in the association between dietary factors and risk of disease. In this study, self-reported dietary assessments were complemented by biomarkers of dietary intake. Dietary and serum measurements of folate and vitamin-B6 from two nested case-control studies within the European Prospective Investigation into Cancer and Nutrition (EPIC) study were integrated in a Bayesian model to explore the measurement error structure of the data, and relate dietary exposures to risk of site-specific cancer. A Bayesian hierarchical model was developed, which included: 1) an exposure model, to define the distribution of unknown true exposure (X); 2) a measurement model, to relate observed assessments, in turn, dietary questionnaires (Q), 24-hour recalls (R) and biomarkers (M) to X measurements; 3) a disease model, to estimate exposures/cancer relationships. The marginal posterior distribution of model parameters was obtained from the joint posterior distribution, using Markov Chain Monte Carlo (MCMC) sampling techniques in JAGS. The study included 554 and 882 case/control pairs for kidney and lung cancer, respectively. In the measurement error component, the error correlation between Q measurements of vitamin-B6 and folate was estimated to be equal to 0.82 (95% CI: 0.76, 0.87). After adjustment for age, center, sex, BMI and smoking status, the kidney cancer odds ratios (OR) were 0.55 (0.16, 1.31) and 1.07 (0.33, 3.44) for one standard deviation increase of vitamin-B6 and folate, respectively. For lung cancer ORs were 0.85 (0.27, 2.42) for vitamin-B6 and 0.55 (0.14, 1.39) for folate. Bayesian models offer powerful solutions to handle complex data structures. After accounting for the role of measurement error, folate and vitamin-B6 were not associated to the risk of kidney and lung cancer.

## 1 Introduction

In nutritional epidemiology, estimating unbiased associations between dietary and lifestyle factors and the risk of chronic diseases presents statistical and epidemiological challenges. One long-standing issue is the presence of measurement errors in self-reported assessments of dietary exposure [9]. In the univariate case, with one error-prone exposure, the power to detect diet-disease associations is reduced by measurement error. Under the classical measurement error model, characterized by random errors with no systematic tendency to under- or over-report intakes, measurement error causes attenuation of relative risk estimates. However, in more common multivariate scenarios with more dietary variables, measurement errors can cause either under- or over-estimation of risk estimates [4].

Due to the importance of measurement error in nutritional epidemiology, statistical models have been developed to improve the estimation of the relationship between diet and disease risk. These models rely on strong conditional independence assumptions. Chief among these is that the measurement errors are non-differential with respect to disease status. This assumption precludes the use of such models in retrospective studies but the non-differential error assumption is well founded in prospective studies when dietary assessments and biological samples for biomarker measurements are taken years before diagnosis. It is also common for models to assume independence of measurement errors between different dietary assessment methods.

Methodology has been developed to allow the potential dependence of the systematic component of measurement errors to be dependent upon individual characteristics such as body mass index, age, and ethnicity [31]. In addition, methods have been proposed to complement self-reported measurements with objective measurements of exposure, but these were primarily implemented in studies evaluating the performance of self-reported assessments and objective measurements of dietary exposure [30, 32, 24] or physical activity [18, 7].

Biomarker measurements of dietary intake have been advocated in two studies using integrated models [25, 8]. In the first study, biomarker measurements of a specific nutrient collected on a sub-sample of the main study were used to calibrate self-reported questionnaire assessments on the same nutrient available on the whole study population [25]. In that work, biomarker levels were assumed to be related to unknown true intake by the classical measurement error model, an assumption unlikely to hold for the vast majority of concentration biomarkers [15]. In the second study, several models were evaluated involving interesting novel relationships between true dietary intake and true biomarker levels, although the measurement error structure of quantities involved was not evaluated [8].

In the current study, we evaluated the relationships between two B-vitamins – folate (vitamin-B9) and vitamin-B6 – and the risk of, in turn, kidney and lung cancer. Data were generated in two nested case-control studies within the European Prospective Investigation into Cancer and Nutrition (EPIC) [27]. Previous analyses of these data showed that plasma levels of Vitamin B6 were inversely associated with kidney cancer [13], and serum levels of Vitamin B6 were inversely related to lung cancer [14]. We analysed serum and plasma vitamin levels, considered as concentration biomarkers (M), along with self-reported dietary questionnaire (Q) and 24-hour dietary recalls (24-HDR, R) as imperfect measurements of B-vitamin intake. Our approach extended previous work using self-reported measurements only [5] in a Bayesian framework [28], and included three components: an exposure, a measurement error, and a disease models. Using data from M measurements, the assumption used in previous work [5], i.e. that errors in Q and R measurements were uncorrelated, was relaxed. The challenge and the limitations of using self-reported and biomarker measurements in a Bayesian framework were illustrated and discussed.

## 2 Statistical Methods

### 2.1 The EPIC data

EPIC is a large scale cohort study aimed at investigating the relationships between lifestyle factors and risk of cancer, and other disease outcome and overall mortality. Its recruitment procedures, collection of questionnaire data, anthropometric measurements, and blood samples have been described in detail elsewhere [27]. In short, information on dietary and other lifestyle factors was collected between 1992 and 2000 from 519,978 study participants in 10 countries across Europe. Blood samples from 385,747 participants were collected at recruitment according to a standardized protocol, and subsequently stored in liquid nitrogen tanks at the International Agency for Research on Cancer (IARC), Lyon, France, at -196^°^C. Information on habitual dietary intakes was assessed at baseline using dietary questionnaire measurements, Q, developed and validated in each participating country [27]. In addition, a single 24-HDR interview, R, was conducted in EPIC to obtain a reference measurement from a subsample (8%) of each cohort [29]. In contrast to Q, R measurements were highly harmonised across countries using the same structure and interview procedure and a common computer program (EPIC-SOFT). Data on anthropometry, physical activity, smoking habits and prevalent chronic conditions were collected using country-specific lifestyle questionnaires.

In this study, the relationship between vitamin-B6 and folate (vitamin-B9) and the risk of kidney and lung cancer was evaluated by modelling serum biomarker M measurements and self-reported Q and R assessments of B-vitamins using data available in two EPIC nested case-control studies [14, 13], and included a total of 2,872 study participants.

The lung cancer study included participants that provided blood samples at recruitment from 8 participating countries: France, Germany, Greece, Italy, Spain, Sweden, the Netherlands, United Kingdom [14]. A total of 882 lung cancer cases were selected on the basis of the *International Classification of Diseases for Oncology, Second Edition* (ICD-O-2), and included all invasive cancers coded as C34. One control per case, matched on country of origin, sex, date of blood collection (±1 month, relaxed to ±5 months for sets without available controls) and date of birth (±1 year, or ±5 years), were selected for the analysis.

In the kidney cancer study, 554 renal cell cancer cases were selected, with the ICD-O-2 code C64.9 [13]. One control was matched to each case according to the same matching criteria as in the lung cancer study. All biochemical analyses were undertaken in the same laboratory under the same conditions, at the same time for all cases and controls in the same batches, and were performed at Bevital A/S (http://www.bevital.no), Bergen, Norway.

### 2.2 The Bayesian hierarchical model

Hierarchical models are used to build complex models through the definition of simpler conditional independence relationships, for which each variable in the model is conditionally related to only a few others. In this way, models are generally characterized by the expression of a marginal model through a sequence of conditional models. The marginal posterior distribution of model parameters is obtained from the joint posterior distribution, using MCMC sampling techniques [10, 21].In this way, complex problems are broken down into modular, possibly hierarchical components that have a relatively simple structure, allowing the complex nature of measurements, in this study dietary and biomarker data, to be accounted for. In extension to the previous work in EPIC using self-reported dietary assessments only [5] and the seminal work of Richardson and Gilks on measurement errors in a Bayesian framework [28], the following model was developed in this study. Let *i* = 1, …, *n* denote study participants, *k* = 1, 2 indicate the two dietary exposures, vitamin-B6 and folate, and *p* = 1, 2 express the disease outcome, in turn kidney and lung cancer. The following notation was used for the various quantities involved in the study:

> *Q*_*ik*_ : dietary questionnaire,
>
> *R*_*ik*_ : 24-hour dietary recall,
>
> *M*_*ik*_ : dietary biomarker measurement,
>
> *Y*_*ip*_ : the disease indicator,
>
> *X*_*ik*_ : true unknown habitual dietary intake.

While Q measurements assessed dietary intake during the year preceding the participants’ recruitment, and were assumed to provide and estimate of habitual intake, R measurements provided estimates of dietary intake during the day preceding the interview. They are labour-intensive and costly to collect, but they are assumed to provide common reference measurements across country-specific dietary data [16]. R measurements were available only on a random subsample of 8% of the study population. These data were considered missing completely at random (MCAR). Biomarkers measurements M were defined as biochemical indicators of short- or medium-term dietary intake [23], and were assumed to provide objective assessments of exposure, as they do not rely on individuals’ ability to recall past dietary exposure. For this reason, they are assumed to be characterized by exposure misclassification with respect to their relationship with the (unknown) true intake, but errors in biomarker measurements are hypothesized to be independent from errors in self-reported dietary assessments [15, 4].

All Q, R and M measurements were log transformed to approximate symmetric distributions. Control of potentially influencing factors was performed using the residual method for R (regressed on age and sex), Q (age, sex and country) and M (age, sex, country, batch, study, and a composite variable expressing smoking status and intensity) measurements. A Bayesian hierarchical model was developed through three structural components [19], which entailed the formulation of an exposure, a measurement error, and a disease model.

#### 2.2.1 The exposure model

True unknown habitual dietary intake (*X*_*ik*_) is not identifiable without making strong assumptions about the unbiasedness of the dietary assessment methods. *X*_*ik*_ represents the latent factor in the hierarchical model. The distribution of true intake was therefore defined on an abstract scale as a bivariate normal distribution *X*_*ik*_ ∼ MVN(0, Σ_*X*_) where

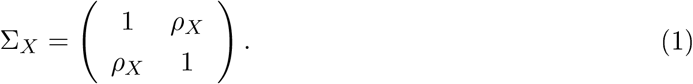

Hence *X*_*i*1_ and *X*_*i*2_ had marginal standard normal distributions and the correlation parameter *ρ*_*X*_ allowed for the possibility that the true intakes of vitamin-B6 and folate were correlated.

#### 2.2.2 The measurement error model

The Q, R and M measurements were related to unknown true intakes by the following linear functional relationships, jointly for vitamin-B6 and folate, as

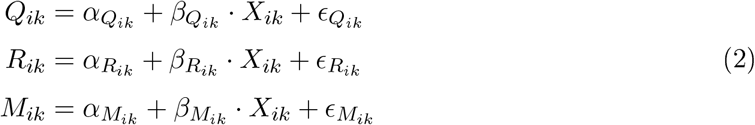

The coefficients *α* and *β* in equation (2) expressed constant and proportional scaling biases, respectively [16], while the *ϵ* terms modelled the random measurement errors in *Q*_*ik*_, *R*_*ik*_ and *M*_*ik*_, and were assumed to be uncorrelated with the true level, *X*_*ik*_ [15, 6].

We assumed that the error terms were unbiased 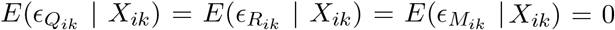 and homoskedastic 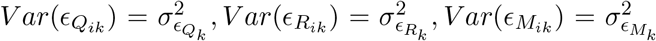. A strong assumption in model (2) was that 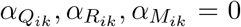, indicating that there were no intercept parameters, since the measurements (*Q, R, M*) were already centered by applying the residual method on them.

Equation (2) also assumes a non-zero error correlations between self-reported dietary measurements, *Q* and *R*, for ∀*k, j* = 1, 2, as also assumed in equation 2, implying

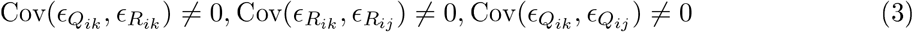

while it was assumed that errors in vitamin-B6 and folate M measurements for were mutually uncorrelated and uncorrelated with errors in Q and R measurements, as

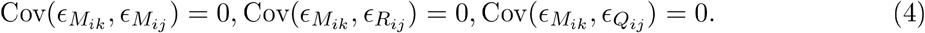

A unique vector of measurements, *Diet* = (*Q*_*ik*_, *R*_*ik*_, *M*_*ik*_), was modelled with the assumption that

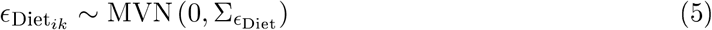

where 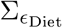 was a 6 × 6 matrix modelled with prior distributions detailed in section 2.3.1. Under the MCAR assumption, missing data in R measurements were handled in MCMC, sampling from the full conditional distribution, given available *Q* and *M* measurements at each iteration. The full conditional distribution can be derived from equations (2) and (5). The facility to sample from partly observed multivariate normal distributions was added as a new feature to JAGS [21] in response to this requirement.

#### 2.2.3 The disease model

The probability *π*_*ip*_ of participant *i* to develop disease *p*, in turn kidney and lung cancer, was related to latent estimates of vitamin-B6 (*X*_*i*1_) and folate (*X*_*i*2_) in analyses that accounted for the study-specific matched design, and adjustment for BMI (continuous) and smoking status (categorical: never, former, current, unknown), denoted by **Z**_*ip*_, as

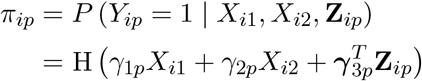

where H indicated the logistic function. The *π*_*ip*_ terms were summed up all individuals in the same case-control stratum 𝒮_*i*_, as

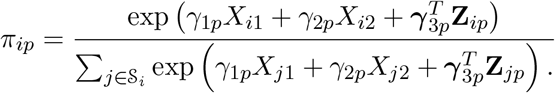

Since *X*_*i*1_ and *X*_*i*2_ had marginal standard deviation of 1 according to equation (1), the parameters *γ*_1_ and *γ*_2_ correspond to a log relative risk for one standard deviation change in true intake since *X* has unitary variance as specified in (1).

### 2.3 Implementation and validation of the Bayesian model

The marginal posterior distribution of model parameters was obtained from the joint posterior distribution, using MCMC [22]. The Bayesian model was implemented in JAGS [21], a program for analysis of Bayesian hierarchical models that can be launched through the R Software [26]. In this work, three chains were run simultaneously using initial values for model parameters that were randomly generated according to normal distributions. After a burn-in of 15,000 iterations and a thinning of 10, a total of 10,000 samples were generated.

#### 2.3.1 Prior distributions

The following prior distributions were assumed for the model parameters:

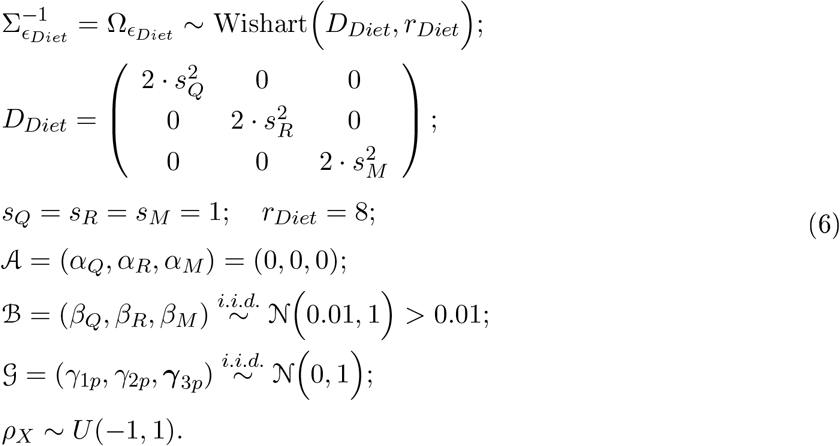

For the precision matrix of the *Diet* vector, representing the error terms for the measurement error model, an unscaled Wishart prior distribution with scale matrix *D*_*Diet*_ and rank *r*_*Diet*_ was used. The scale matrix has dimension 6 × 6 = *m* × *m* due to the bivariate (vitamin– B6 and folate) measurements for (*Q, R, M*). It is characterized by a diagonal matrix, were the off-diagonal element are null, and the main ones are described by twice the square of a scale parameter *s*_*Q*_ = *s*_*R*_ = *s*_*M*_ = 1. The importance of the scale parameters for the Wishart prior distribution, that achieve arbitrarily high noninformativity of all standard deviation and correlation parameters, is described in [11]. While the rank of the Wishart distribution is set to 8 since the rule of thumb is the *m* + 1, however in this case was bumped up by 1 to penalize error correlations near [1, −1].

The vectors 𝒜 and ℬ refer respectively to the constant and proportional scaling biases. The first one, 𝒜, is assigned to zero, since residuals data are used. While for the second one, ℬ, each of the *β* priors parameters is bounded away from zero (ℬ *>* 0.01) to avoid a singularity (i.e. model collapses so that all observed variation is explained by measurement error). This is consistent and possible with our prior beliefs, 𝒩 (0.01, 1), i.e. we believe that these measurements have a non-zero correlation with true intake. Sensitivity analyses were performed to choose the right cut-off point such that no singularity and/or exclusion of plausible values occurs. The vectors 𝒢 refer to the *γ* parameters in the disease model, respectively, for the “true” intake and the further covariates. The priors for each element in 𝒢 are weakly informative with a standard normal distribution 𝒩 (0.01, 1).

The prior distribution for “true” intake *X* is bivariate normal with mean zero, unit standard deviation and correlation *ρ*, as described in (1). The true intake is unidentifiable so we define it on an arbitrary scale where its mean and standard deviation are fixed. For this reason, we set an uniform distribution within the interval (1, −1) on *ρ*_*X*_, the correlation between true intakes of vitamin-B6 and folate.

The choice of the prior distribution is crucial. Initially we used the scaled Inverse-Wishart distribution as a prior for the variance-covariance matrix of the errors. Assigning hyper-parameters, i.e. prior scale and degrees of freedom, is straightforward, as they can be interpreted in terms of the standard deviation of each error component [11].

Assigning hyper-parameters, i.e. prior scale and degrees of freedom, was straightforward, given the conjugate family of prior distributions. As shown in [11], members of the family possess the attractive property of all standard deviation and correlation parameters being marginally non-informative for particular hyper-parameter choices.

However, it was not possible to use the Inverse-Wishart distribution for our measurement-error model due to identifiability, which led to over-parameterization and to an unidentifiable model. Also, a scaled Wishart prior distribution resulted in error variances for any of the measurements to collapse to zero, causing mixing problems. Therefore, a standard Wishart distribution allowed the identifiability and the mixing problems to be solved. A non-informative standard Wishart prior distribution for the elements of the variance-covariance matrix was initially used, and then we moved to a more informative standard Wishart prior distribution to have more stable estimates. The hyper-parameters of the Wishart were chosen by conducting sensitivity analyses, where results were checked in relation to model assumptions, including the linearity of the relationships in the measurement error model and the hypothesis of reference measurement for the 24-hour dietary recall, R.

The final hyper-parameters of the standard Wishart, as in (2.3.1), were chosen by applying the model assumptions (3) and (4) in the symmetry of the variance-covariance matrix and in the choice of null values for the off-diagonal elements. Sensitivity analyses were conducted, with small perturbations of the initial values, to check that results verified the other model assumptions of the measurement error model (2): the linearity and the reference measurement for the 24-hour dietary recall, R.

#### 2.3.2 Frequentist analysis

In order to evaluate the identifiability of model parameters, moment estimators were derived from the structural equations formulation for specific quantities of the exposure and measurement error model components (Supplementary Material). Given the linear associations in model (2) and the related assumptions on error variances and covariances, the population covariance matrix (Σ) of a set of observed variables (Q, R, M) is a function of a set of parameters *q* = (*q*_1_, …, *q*_*p*_), as 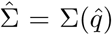 [1, 2]. In particular, the variance of true exposure was estimated as a function of the covariance between the biomarkers of the vitamin-B6 and folate, i.e. Cov(*M*_*i*1_, *M*_*i*2_), and the covariance between biomarker and 24-HDR measurements, i.e. Cov(*M*_*i*1_, *R*_*i*1_) and Cov(*M*_*i*1_, *R*_*i*2_). The terms were usually very weakly related to each other, especially in the case of one replicate of R measurements, due to the large within-person variability, as in our study. This observation calls for the availability of replicates of R measurements in future studies to make data most informative [20]. Conditional logistic regression models were used to estimate the odds ratio (OR) and associated 95% confidence intervals (CI) of, in turn, kidney and lung cancer in relation to Q and M measurements of vitamin-B6 and folate. Models were minimally adjusted by matching factors, and also fully adjusted by BMI and smoking status. The comparison with RR estimates from the Bayesian model allowed the extent of measurement error correction to be evaluated. In line with Freedman and colleagues [8], principal component scores obtained from Q, R and M measurements of, in turn, vitamin-B6 and folate were related to the risk of kidney and lung cancers in conditional logistic models. All analyses were performed in R [26] and JAGS [21].

## 3 Results

The characteristics of the study population of the kidney and lung cancer nested case-control studies are shown in Table 1. Our study included 554 and 882 case-control pairs in the kidney and lung cancer studies, respectively. The correlation coefficients between Q, R and M measurements of vitamin-B6 and folate are displayed in Table 2. Values indicate fairly weak level of agreement overall, not only between biomarker and self-reported measurements (consistently below 0.15), but also between Q and R measurements (consistently below 0.25).

**Table 1:**
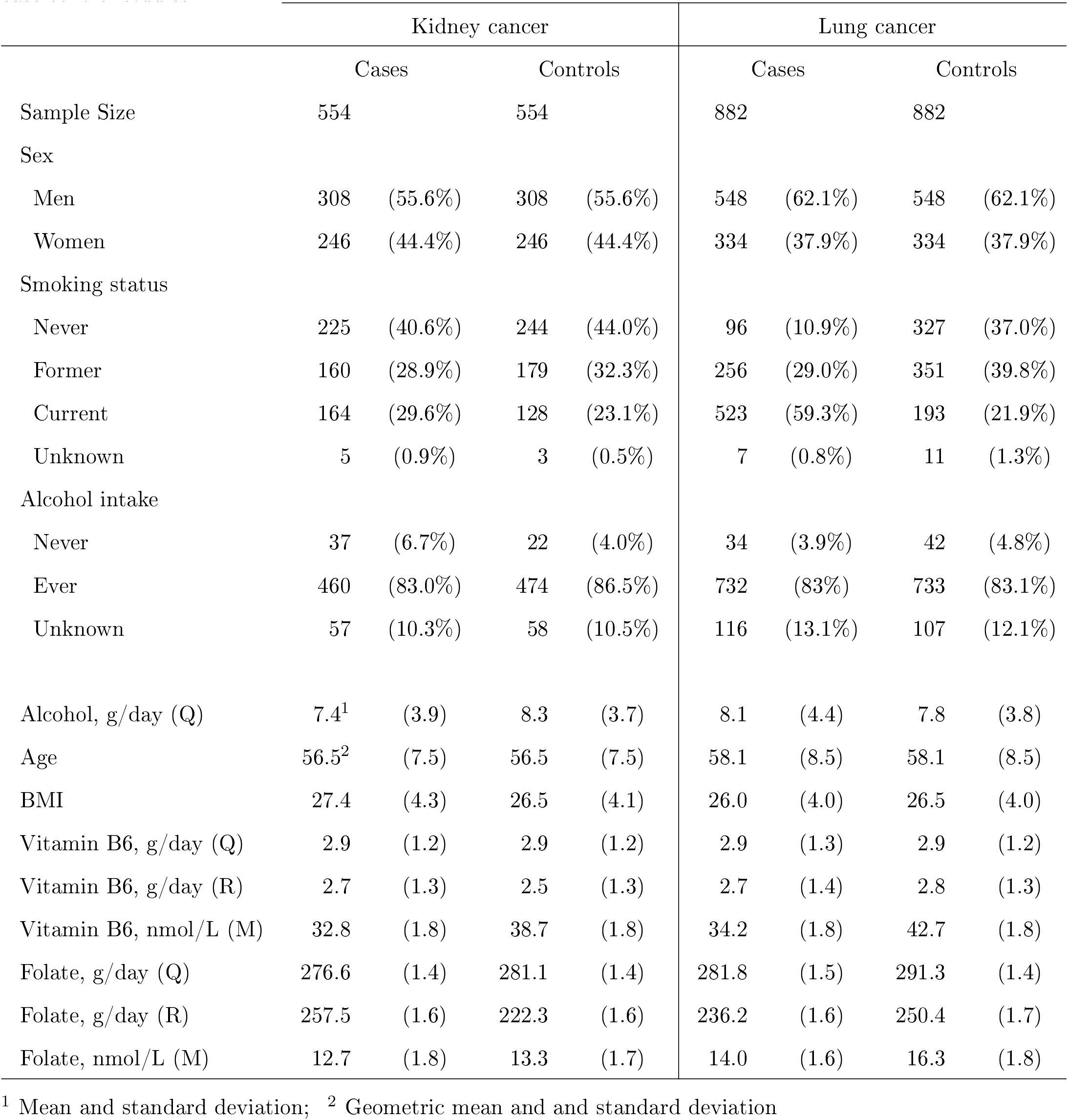
Characteristics of the study population in the EPIC kidney and lung cancer nested case-control studies.

|  | Kidney cancer |  |  |  | Lung cancer |  |  |  |
| --- | --- | --- | --- | --- | --- | --- | --- | --- |
|  | Cases |  | Controls |  | Cases |  | Controls |  |
| Sample Size | 554 |  | 554 |  | 882 |  | 882 |  |
| Sex |  |  |  |  |  |  |  |  |
| Men | 308 | (55.6%) | 308 | (55.6%) | 548 | (62.1%) | 548 | (62.1%) |
| Women | 246 | (44.4%) | 246 | (44.4%) | 334 | (37.9%) | 334 | (37.9%) |
| Smoking status |  |  |  |  |  |  |  |  |
| Never | 225 | (40.6%) | 244 | (44.0%) | 96 | (10.9%) | 327 | (37.0%) |
| Former | 160 | (28.9%) | 179 | (32.3%) | 256 | (29.0%) | 351 | (39.8%) |
| Current | 164 | (29.6%) | 128 | (23.1%) | 523 | (59.3%) | 193 | (21.9%) |
| Unknown | 5 | (0.9%) | 3 | (0.5%) | 7 | (0.8%) | 11 | (1.3%) |
| Alcohol intake |  |  |  |  |  |  |  |  |
| Never | 37 | (6.7%) | 22 | (4.0%) | 34 | (3.9%) | 42 | (4.8%) |
| Ever | 460 | (83.0%) | 474 | (86.5%) | 732 | (83%) | 733 | (83.1%) |
| Unknown | 57 | (10.3%) | 58 | (10.5%) | 116 | (13.1%) | 107 | (12.1%) |
| Alcohol, g/day (Q) | 7.4<sup>1</sup> | (3.9) | 8.3 | (3.7) | 8.1 | (4.4) | 7.8 | (3.8) |
| Age | 56.5<sup>2</sup> | (7.5) | 56.5 | (7.5) | 58.1 | (8.5) | 58.1 | (8.5) |
| BMI | 27.4 | (4.3) | 26.5 | (4.1) | 26.0 | (4.0) | 26.5 | (4.0) |
| Vitamin B6, g/day (Q) | 2.9 | (1.2) | 2.9 | (1.2) | 2.9 | (1.3) | 2.9 | (1.2) |
| Vitamin B6, g/day (R) | 2.7 | (1.3) | 2.5 | (1.3) | 2.7 | (1.4) | 2.8 | (1.3) |
| Vitamin B6, nmol/L (M) | 32.8 | (1.8) | 38.7 | (1.8) | 34.2 | (1.8) | 42.7 | (1.8) |
| Folate, g/day (Q) | 276.6 | (1.4) | 281.1 | (1.4) | 281.8 | (1.5) | 291.3 | (1.4) |
| Folate, g/day (R) | 257.5 | (1.6) | 222.3 | (1.6) | 236.2 | (1.6) | 250.4 | (1.7) |
| Folate, nmol/L (M) | 12.7 | (1.8) | 13.3 | (1.7) | 14.0 | (1.6) | 16.3 | (1.8) |

<sup>1</sup> Mean and standard deviation; <sup>2</sup> Geometric mean and standard deviation

**Table 2:** Observed correlation coefficients, 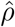, between Q, R and M measurements for vitamin-B6 and folate intakes.

|  |  | Vitamin-B6 |  |  | Folate |  |  |
| --- | --- | --- | --- | --- | --- | --- | --- |
| | | $Q_1$ | $R_1$ | $M_1$ | $Q_2$ | $R_2$ | $M_2$ |
| Vitamin-B6 | $Q_1$ | 1 | | | | | |
| | $R_1$ | 0.222 | 1 | | | | |
| | $M_1$ | 0.132 | 0.113 | 1 | | | |
| Folate | $Q_2$ | 0.856 | 0.180 | 0.072 | 1 | | |
| | $R_2$ | 0.182 | 0.644 | 0.150 | 0.221 | 1 | |
| | $M_2$ | 0.100 | 0.087 | 0.319 | 0.101 | 0.194 | 1 |

Estimates from the exposure model are reported in Table 3, for the observed measurements (i.e., Q, R and M) and the latent factors (X) of vitamin-B6 and folate. After log-transformation and computation of residuals, the standard deviations (*σ*) were larger for R measurements, 0.452 and 0.487, than Q measurements, 0.290 and 0.314, for vitamin-B6 and folate measurements respectively, likely reflecting large within-person variability of one replicate of R measurements compared to Q assessments. The standard deviation of M measurements was 0.594 for vitamin-B6 and 0.530 for folate.

**Table 3:**
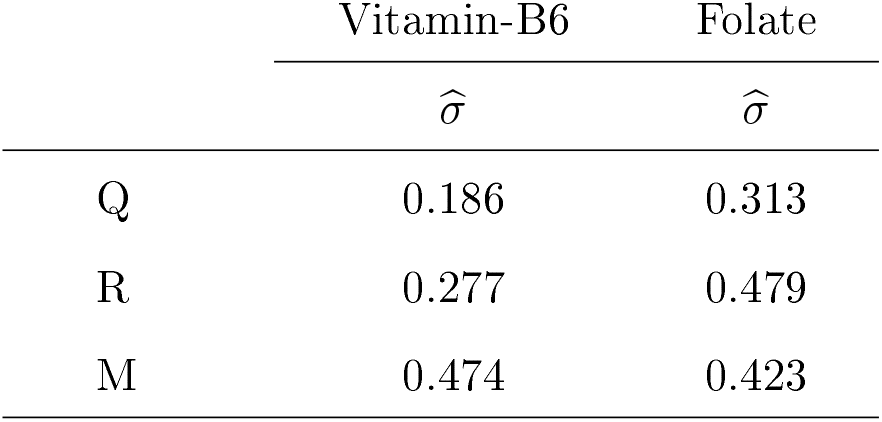
Observed standard deviation terms, 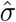, between Q, R and M measurements for vitamin-B6 and folate intakes.

|  | Vitamin-B6 | Folate |
| --- | --- | --- |
| | $\hat{\sigma}$ | $\hat{\sigma}$ |
| Q | 0.186 | 0.313 |
| R | 0.277 | 0.479 |
| M | 0.474 | 0.423 |

Estimates of the measurement error model are presented in Table 4. For vitamin-B6 error terms are particularly large for M measurements (0.213, 95%CI: 0.182–0.242) than R (0.065, 95%CI 0.054–0.078) and Q (0.033, 95%CI 0.031–0.035) measurements. Error terms for folate intake are more comparable across the three types of assessment. Error correlations between vitamin-B6 and folate were particularly large for Q than R measurements, while independence of errors was assumed between the two biomarkers.

**Table 4:** Measurement error model: estimated error variance 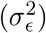 and correlation (*ρ*_*ϵ*_) terms for Q, R and M measurements.

| | Vitamin-B6 | | Folate | | $\hat{\rho}_\epsilon$ |
| --- | --- | --- | --- | --- | --- |
| | $\hat{\sigma}_\epsilon^2$ | (95% CI<sup>1</sup>) | $\hat{\sigma}_\epsilon^2$ | (95% CI<sup>1</sup>) | |
| Q | 0.035 | (0.033, 0.037) | 0.098 | (0.092, 0.103) | 0.804 |
| R | 0.077 | (0.064, 0.091) | 0.229 | (0.181, 0.278) | 0.589 |
| M | 0.224 | (0.126, 0.305) | 0.179 | (0.122, 0.222) | 0 |

<sup>1</sup>Credible Intervals

Parameter estimates obtained in the disease model are shown in Tables 5 and 6. Estimates express, in turn, kidney and lung cancer relative risk (RR) associated to a change of one standard deviation (1-SD) of vitamin-B6 and folate Q and M measurements, and the latent factors X. For kidney cancer, no association was observed for Q measurements, while M measurements of vitamin-B6 were inversely related to the risk, with RR equal to 0.77 (95% CI: 0.67, 0.88). For lung cancer and lung cancer (0.77, 95%CI 0.67–0.88), and M measurements of folate were inversely related to the risk of lung cancer (). After measurement error correction, and adjustment for BMI and smoking history, X measurements of vitamin-B6 and folate intakes were not associated with the risk of, in turn, kidney and lung cancer.

**Table 5:** Disease model: relative risk 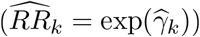 estimates for kidney cancer associated to a 1-SD increase in exposure.

|  | Vitamin-B6 |  | Folate |  |
| --- | --- | --- | --- | --- |
| | $\widehat{RR}_1$ | (95% CI<sup>1</sup>) | $\widehat{RR}_2$ | (95% CI<sup>1</sup>) |
| Q | 1.08 | (0.84, 1.38) | 0.90 | (0.71, 1.15) |
| R | - | - | - | - |
| M | 0.77 | (0.67, 0.89) | 0.99 | (0.84, 1.07) |
| X | 0.59 | (0.21, 1.15) | 1.01 | (0.47, 2.66) |

<sup>1</sup>Credible Intervals; $X^*$ = Principal component scores

**Table 6:** Disease model: relative risk 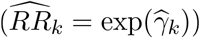 estimates for lung cancer associated to a 1-SD increase in exposure.

|  | Vitamin-B6 |  | Folate |  |
| --- | --- | --- | --- | --- |
| | $\widehat{RR}_1$ | (95% CI<sup>1</sup>) | $\widehat{RR}_2$ | (95% CI<sup>1</sup>) |
| Q | 1.03 | (0.81, 1.32) | 0.93 | (0.81, 1.32) |
| R | - | - | - | - |
| M | 0.76 | (0.67, 0.88) | 0.87 | (0.76, 0.99) |
| X | 0.71 | (0.21, 2.33) | 0.42 | (0.12, 1.25) |

The principal component scores of vitamin-B6 were borderline inversely related to the risk of kidney cancer (RR = 0.88, 95% CI: 0.77, 1.00) and lung cancer (RR = 0.88, 95% CI: 0.78, 1.00), while no association was observed for folate.

## 4 Discussion

In epidemiological studies evaluating the relationship between disease risk and exposure affected by measurement errors, Bayesian models offer flexible opportunities to handle complex data structure, for example integrating information from self-reported assessments and biomarker concentrating levels together with disease indicators, thus combining features of validation and risk models in a single analytical framework [28]. In the EPIC study, a Bayesian latent factor hierarchical model extended previous work [5] to evaluate the relationships between vitamin-B6 and folate intakes with risk of, in turn, kidney and lung cancers. Within the model, the measurement error structure of all observed quantities was investigated, including error correlations in self-reported assessments, and the variability of true intakes was estimated. The model also included an etiological component to estimate the association between dietary exposures and risk of disease.

Dietary biomarkers have become increasingly popular because they can provide information about nutrients’ bioavailability, defined as the available effective internal dose after absorption and metabolism [12]. Two classes of dietary biomarkers can be distinguished. First, biomarker based on the metabolic balance between intake and excretion of specific chemical components, i.e. the percent recovery of the compound or its metabolites in excretion products, mostly in urinary samples, over a fixed period of time [17]. They are defined *recovery* biomarkers, and they provide estimates of absolute intakes. It is customarily assumed that the classical measurement error model applies, as *M*_*i*_ = *T*_*i*_ + *ϵ*_*M*_. The list of *recovery* biomarkers is very limited, i.e. the doubly labelled water collected in urines to estimate absolute levels of total energy intake, urinary nitrogen and urinary potassium for dietary protein and potassium, respectively [12]. The second class of biomarkers are measured as concentration of specific compounds in biological fluids [17], and are known as *concentration* markers. They do not have the same quantitative relationship with dietary intake levels for each study participants, and they cannot be translated into absolute levels of intake [12].

In this study, *concentration* biomarkers were modelled with dietary assessments, thus allowing for components of random (*ϵ*_*M*_) systematic errors (*α*_*M*_ and *β*_*M*_, constant and proportional scaling bias, respectively) in Model (2) [15]. These terms capture analytical variation and technical errors. The combined use of self-reported dietary assessments and objective biomarker measurements in etiological models has been the object of research. Freedman and colleagues developed relevant work hypothesizing a relationship between true dietary intake (TDI) as a determinant of true biomarker level (TBL) [8]. Reported dietary intake and measured (concentration or recovery) biomarker levels were linearly related, respectively, to TDI and TBL, under the assumption of independent and non-differential (to disease status) error structure. The Authors advocate the use of feeding studies to quantify the relationship between TDI and TBL. In the same study, the use of principal component analysis for self-reported and biomarker measurements was suggested as a good strategy to model biomarker and dietary data jointly. In our work, the principal component score of vitamin-B6 provided an inverse borderline statistically significant association for, in turn, kidney and lung cancer.

Our study had several limitations. First, linear relationships in the measurement error model were linking self-reported and objective (log-transformed) measurements to unknown true intake in the equations of measurement error model (2). Second, the error model relied on the assumption that R provided reference measurements, an assumption that was shown to be weak in validation studies that used *recovery* biomarkers as reference assessments ([18]). Nevertheless our model included terms to incorporate components of systematic errors such as error correlation between self-reported assessments. Third, R measurements were available by design only in the 8% of the study population [29], i.e. for study participants that were part of the EPIC calibration study. Although MCMC was technically handling missing data, this represents a sizeable loss of information. Also, only one replicate of R measurements per participant was available, thus resulting in large variability attributable to within-person variation. Future applications on the model developed in this study should include more informative sets of data, with several replicates of the reference measurements R, possibly available on the totality of study participants.

Within this study, the time distance between the assessment of dietary measurements and the collection of biological samples was evaluated. The consistency of correlation values between Q and R measurements were evaluated, when dietary and biological measurements were collected within one month, 6 months or within one year. Overall this evaluation showed marginal impact of time on the stability of correlation estimates, a results at least partly attributable to the nature of dietary components investigated in this work. It is likely that the intakes of dietary sources of vitamin-B6 and folate were fairly stable within study participants’ habitual diet.

Overall, the results of this study suggested that vitamin-B6 and folate were not associated to the risk of, in turn, kidney and lung cancer, after accounting for the error structure of all measurements involved in the estimation process. This evidence was not consistent with previous observations reporting that biomarker measurements of these B-vitamins were associated to the risk of developing kidney [13] and lung cancer [14]. Other than the limitations already discussed, the lack of relationships assessed in this study could be due to the particularly challenging setting of this study. Despite self-reported dietary and blood measurements of vitamin-B6 and folate are supposed to quantify the same intake, in practice they estimate different dietary quantities. As a result, by imposing a common relational structure among Q, R and M measurements around unknown true intake, the level of uncertainty outweighs the strength of the signal in the relationship between exposure and risk of cancer. In line with [8], future extensions of our work could relax the assumption of a unique common latent structure for Q, R and M measurements, and rather adopt a more realistic model, where TDI is related to Q, and TBL is related to M through the classical measurement error model. External information on the link between TDI and TBL from feeding or functional studies could be fairly easily incorporated in the model.

Bayesian modelling lends itself as a powerful, and its implementation requires specific knowledge due to the complex mathematical theory. In the last 30 years, MCMC sampling techniques have transformed Bayesian statistical inference, allowing Bayesian statistical methods to become widely used for data analysis and modelling [10]. MCMC allows approximate Bayesian inference by drawing a sequence of random samples from the posterior distribution of the parameters, the target of Bayesian inference. It is a computationally intensive method, but the growth in computing power has led its routine application to increasingly large and complex problems. At the same time, continuing theoretical developments have produced sophisticated and efficient sampling methods. In this study, the computing time was 6 hours for one Bayesian model to run with a PC Intel(R) Core i5, CPU 2.40GHz, 8 GB RAM. The model had three chains, a burn-in of 15,000 iterations, an adaptation of 10,000, and final estimates were based on 20,000, with thin equal to 10. *PF: Martyn, how do these numbers look like? Move some of these details to the Methods?* The JAGS package [21] was used in this study, and the R package coda [22] helped evaluate the mixing of MCMC chains and the convergence of posterior distributions. Both visual and statistical criteria were used with the Gelman and Geweke diagnostics [3].

This project triggered some relevant developmental work in the JAGS software, some of which will be incorporated into future releases. As JAGS is general-purpose software for Bayesian modelling and is freely distributed under the terms of the GNU General Public License (GPL), these advances will be available to implement future use of this methodology with other dietary and biomarker variables, possibly displaying larger levels of agreement across different types of assessment, specifically and primarily in terms of Pearson correlation coefficients. In this respect, the growing interest towards biomarkery discovery, for example using metabolomics data (PMID: 28122782), could create novel opportunities to apply this model new sets of data.

## Data Availability

All data produced in the present study is part of the European Prospective Investigation into Cancer and Nutrition (EPIC) study. The EPIC study is one of the largest cohort studies in the world, with more than half a million (521 000) participants recruited across 10 European countries and followed for almost 15 years.

https://epic.iarc.fr/

## Acknowledgements

We thank the World Cancer Research Funds (WCRF International) that supported this study (Grant IARC-2012-10-10-03). We also thank the EPIC PIs to grant their permission for use make secondary use of B-vitamins data in relation to kidney and lung cancers to illustrate the Bayesian model. A special thank for Dr. George Luta (Georgetown University, D.C., USA) for insightful comments to various versions of this manuscript.

## Conflict of interest

No conflict of interest.

## Appendix A

Specifically, the bivariate model equations for the measurement error model reported in detail are:

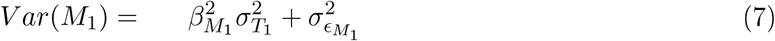

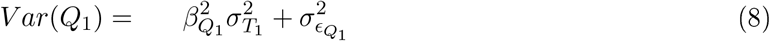

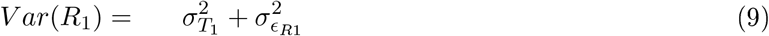

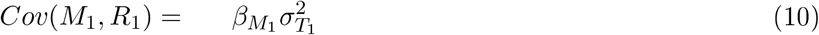

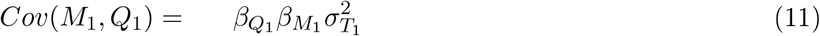

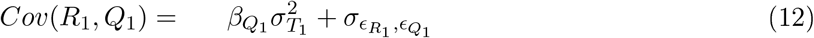

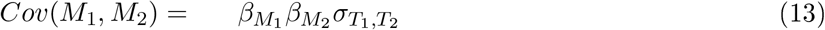

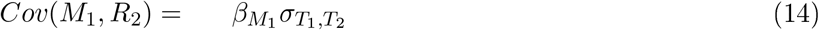

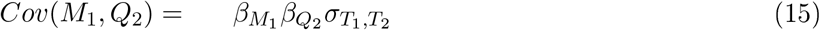

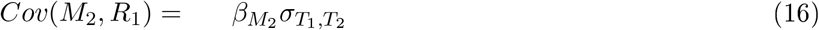

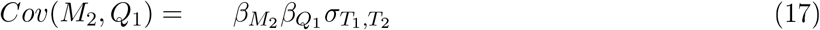

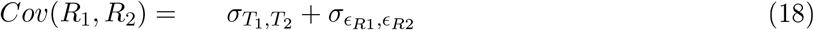

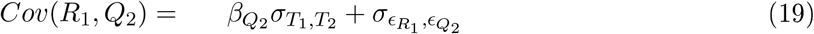

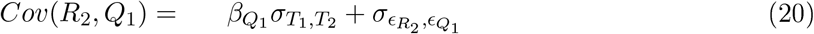

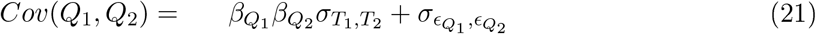

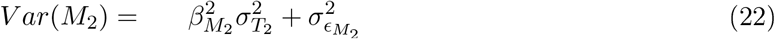

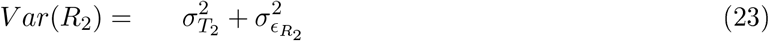

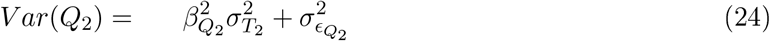

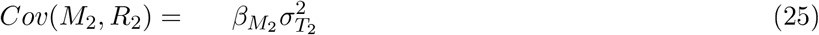

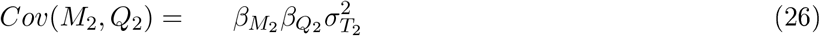

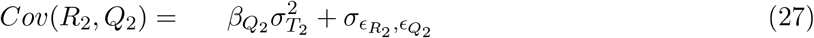

We have, in total, 21 equations and 19 parameters to estimate: 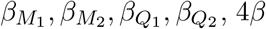 parameters;

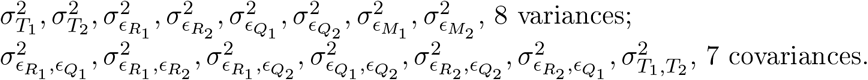

From (13) and (16), we have

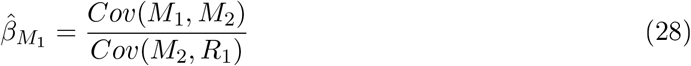

From (13) and (14), we have

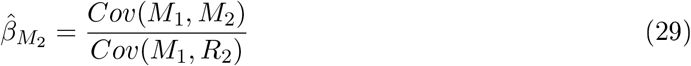

From (10) and (28), we get:

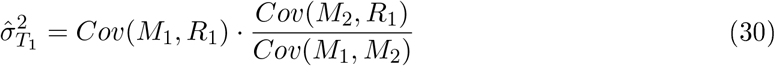

From (25) and (29), we get:

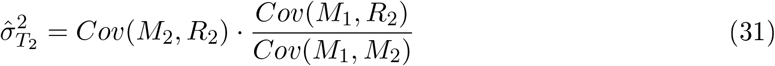

From the combinations of (10) and (11) with (17) and (16), we get:

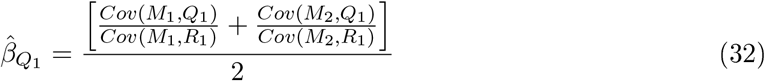

From the combinations of (14) and (15) with (25) and (26), we get:

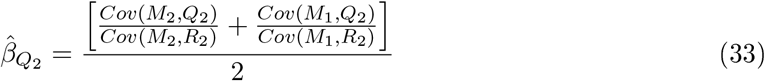

Using the expressions of (28), (29), (32) and (33), we get the covariance estimate between the two intakes. Hence, starting from (14), (17), (13), (15), (16):

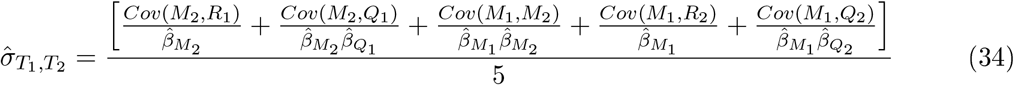

Moreover, in a similar manner as done before, we get the remaining 6 variance estimates related to *M*_*k*_, *R*_*k*_ and *Q*_*k*_, where *k* = 1, 2.

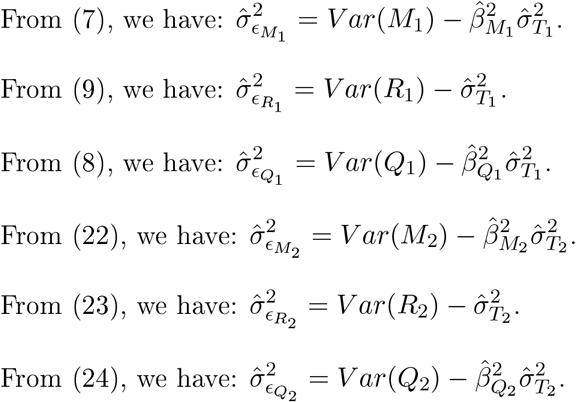

We individuated all the 4*β* parameters, all the 8 variance estimates in total and one covariance parameter. Only 6 estimates related to the food frequency questionnaires and the 24-hours recall are missing. Let’s try to individuate them.

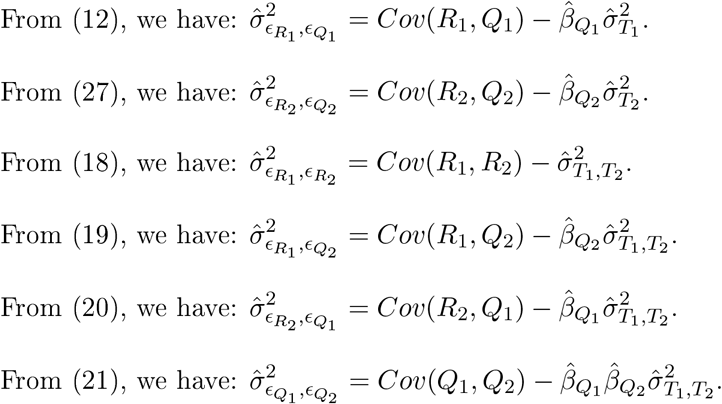

All the 19 parameters have been identified. The bivariate measurement error model with the aforementioned assumptions is identifiable.

## Notes

### Competing Interest Statement

The authors have declared no competing interest.

### Author Declarations

This work was funded by a WCRF International award (IARC-2012-10-10-03). All data produced in the present study is part of the European Prospective Investigation into Cancer and Nutrition (EPIC) study. The coordination of EPIC is financially supported by International Agency for Research on Cancer (IARC) and also by the Department of Epidemiology and Biostatistics, School of Public Health, Imperial College London which has additional infrastructure support provided by the NIHR Imperial Biomedical Research Centre (BRC). National cohorts are supported by local funding Agencies.

## References

[1] P.M. Bentler and D.G. Weeks. Linear structural equations with latent variables. Psychometrika, 45(3):289–308, 1980.

[2] K.A. Bollen. A new incremental fit index for general structural equation models. Sociological Methods & Research, 17(3):303–316, 1989.

[3] M.K. Cowles and B.P. Carlin. Markov chain monte carlo convergence diagnostics: A comparative review. Journal of the American Statistical Association, 91(434):883– 904, 1996.

[4] N.E. Day and P. Ferrari. Some methodological issues in nutritional epidemiology. IARC Scientific Publications, 156:5–10, 2002.

[5] P. Ferrari, R.J. Carroll, P. Gustafson, and E. Riboli. A bayesian multilevel model for estimating the diet/disease relationship in a multicenter study with exposures measured with error: The epic study. Statistics in Medicine, 27(29):6037–6054, 2008.

[6] P. Ferrari, N.E. Day, H.C. Boshuizen, A. Roddam, K. Hoffmann, A. Thiebaut, and G. et al. Pera. The evaluation of the diet/disease relation in the epic study: considerations for the calibration and the disease models. International Journal of Epidemiology, 37(2):368–378, 2008.

[7] P. Ferrari, C. Friedenreich, and C.E. Matthews. The role of measurement error in estimating levels of physical activity. American Journal of Epidemiology, 166(7):832–840, 2007.

[8] L.S. Freedman, V. Kipnis, A. Schatzkin, N. Tasevska, and N. Potischman. Can we use biomarkers in combination with self-reports to strengthen the analysis of nutritional epidemiologic studies? Epidemiologic Perspectives & Innovations, 7(1):2, 2010.

[9] J.L. Freudenheim and J.R. Marshall. The problem of profound mismeasurement and the power of epidemiological studies of diet and cancer. Nutrition and Cancer, 11(4):243–250, 1988.

[10] A. Gelman, J.B. Carlin, H.S. Stern, D.B. Dunson, A. Vehtari, and D.B. Rubin. Bayesian Data Analysis. Chapman&Hall/CRC ed., 2003.

[11] A. Huang and M.P. Wand. Simple marginally noninformative prior distributions for covariance matrices. Bayesian Analysis, 8(2):439 –451, 2013.

[12] M. Jenab, N. Slimani, M. Bictash, P. Ferrari, and S.A. Bingham. Biomarkers in nutritional epidemiology: applications, needs and new horizons. Human Genetics, 125(5-6):507–525, 2009.

[13] M. Johansson, A. Fanidi, D.C. Muller, J.K. Bassett, O. Midttun, and S.E. et al. Vollset. Circulating biomarkers of one-carbon metabolism in relation to renal cell carcinoma incidence and survival. Journal of the National Cancer Institute, 106(12), 2014.

[14] M. Johansson, C. Relton, P.M. Ueland, S.E. Vollset, O. Midttun, O. Nygard, and N. et al. Slimani. Serum b vitamin levels and risk of lung cancer. Journal of the American Medical Association, 303(23):2377–2385, 2010.

[15] R. Kaaks, P. Ferrari, A. Ciampi, M. Plummer, and E. Riboli. Uses and limitations of statistical accounting for random error correlations, in the validation of dietary questionnaire assessments. Public Health Nutrition, 5(6A): 969–976, 2002.

[16] R. Kaaks and E. Riboli. Validation and calibration of dietary intake measurements in the epic project: Methodological considerations. International Journal of Epidemiology, 26(1):S15–S25, 1997.

[17] R. Kaaks, E. Riboli, and R. Sinha. Biochemical markers of dietary intake. IARC Scientific Publications, 1997.

[18] V. Kipnis, A.F. Subar, D. Midthune, L.S. Freedman, R. Ballard-Barbash, R.P. Troiano, S. Bingham, D.A. Schoeller, A. Schatzkin, and R.J. Carroll. Structure of dietary measurement error: Results of the open biomarker study. American Journal of Epidemiology, 158(1):14–21, 2003.

[19] M. Pittavino, A. Dreyfus, C. Heuer, J. Benschop, P. Wilson, J. Collins-Emerson, P. R. Torgerson, and R. Furrer. Comparison between generalized linear modelling and additive bayesian network; identification of factors associated with the incidence of antibodies against leptospira interrogans sv pomona in meat workers in new zealand. Acta Tropica, 173:191–199, 2017.

[20] M. Pittavino, A. Dreyfus, C. Heuer, J. Benschop, P. Wilson, J. Collins-Emerson, P. R. Torgerson, and R. Furrer. Data on leptospira interrogans sv pomona infection in meat workers in new zealand. Data in Brief, 13:587–596, 2017.

[21] M. Plummer. Jags: a program for analysis of Bayesian graphical models using gibbs sampling. Proc 3rd Int Work Dist Stat Comp, 1(20-22):1–10, 2003.

[22] M. Plummer, N. Best, K. Cowles, and K. Vines. Coda: convergence diagnosis and output analysis for mcmc. R News, 6(1):7–11, 2006.

[23] N. Potischman and J.L. Freudenheim. Biomarkers of nutritional exposure and nutritional status: An overview. Journal of Nutrition, 133(3):873S–874S, 2003.

[24] S.R. Preis, D. Spiegelman, B. Zhao, and W. Willett. Random error in biomarkers and the appearance of correlated error between dietary intake measurements. American Journal of Epidemiology, 167(11, S):S87, 2008.

[25] R.L. Prentice, E. Sugar, C. Wang, M. Neuhouser, and R. Patterson. Research strategies and the use of nutrient biomarkers in studies of diet and chronic disease. Public Health Nutrition, 5(6A): 977– 984, 2002.

[26] R Core Team. R: A language and environment for statistical computing. R Foundation for Statistical Computing, Vienna, Austria, 2019.

[27] E. Riboli, K.J. Hunt, N. Slimani, P. Ferrari, T. Norat, M. Fahey, U.R. Charrondiere, B. Hemon, C. Casagrande, J. Vignat, K. Overvad, A. Tjonneland, F. Clavel-Chapelon, A. Thiebaut, J. Wahrendorf, H. Boeing, D. Trichopoulos, A. Trichopoulou, P. Vineis, D. Palli, H.B. Bueno-de Mesquita, P.H.M. Peeters, E. Lund, D. Engeset, C.A. Gonzalez, A. Barricarte, G. Berglund, G. Hallmans, N.E. Day, T.J. Key, R. Kaaks, and R. Saracci. European prospective investigation into cancer and nutrition (epic): study populations and data collection. Public Health Nutrition, 5(6B, SI):1113–1124, 2002.

[28] S. Richardson and W.R. Gilks. Conditional independence models for epidemiological studies with covariate measurement error. Statistics in Medicine, 12(18):1703–1722, 1993.

[29] N. Slimani, R. Kaaks, P. Ferrari, C. Casagrande, F. Clavel-Chapelon, G. Lotze, A. Kroke, D. Trichopoulos, A. Trichopoulou, C. Lauria, M. Bellegotti, M.C. Ocke, P.H.M. Peeters, D. Engeset, E. Lund, A. Agudo, N. Larranaga, I. Mattisson, C. Andren, I. Johansson, G. Davey, A.A. Welch, K. Overvad, A. Tjonneland, W.A van Staveren, R. Saracci, and E. Riboli. European prospective investigation into cancer and nutrition (epic) calibration study: rationale, design and population characteristics. Public Health Nutrition, 5(6B, SI):1125–1145, 2002.

[30] D. Spiegelman, S. Schneeweiss, and A. McDermott. Measurement error correction for logistic regression models with an ‘’alloyed gold standard”. American Journal of Epidemiology, 145(2):184–196, 1997.

[31] E.A. Sugar, C. Y. Wang, and R.L. Prentice. Logistic regression with exposure biomarkers and flexible measurement error. Biometrics, 63(1):143–151, 2007.

[32] D. Thurigen, D. Spiegelman, M. Blettner, C. Heuer, and H. Brenner. Measurement error correction using validation data: a review of methods and their applicability in case-control studies. Statistical Methods in Medical Research, 9(5):447–474, 2000.

